# Geographic Information System Protocol for Mapping Areas Targeted for Mosquito Control in North Carolina

**DOI:** 10.1101/2022.11.14.22282322

**Authors:** Andrew Mueller, Anthony Thomas, Jeff Brown, Abram Young, Kim Smith, Roxanne Connelly, Stephanie L. Richards

**Affiliations:** Brunswick County Geographic Information Systems, 30 Government Center Dr., Bolivia, NC; Brunswick County Mosquito Control, 179 March 9, 1764, Bolivia, NC; Columbus County Health Department, Whiteville, NC; Centers for Disease Control and Prevention, Division of Vector-borne Diseases, Arboviral Diseases Branch, Fort Collins, CO; Environmental Health Sciences Program, Department of Health Education and Promotion, College of Health and Human Performance, East Carolina University, Greenville, NC

**Keywords:** geographic information science, emergency mosquito control, hurricane

## Abstract

Geographic information systems (GIS) can be used to map mosquito larval and adult habitats and human populations at risk for mosquito exposure and possible arbovirus transmission. GIS can help simplify and target mosquito control efforts during routine surveillance and post-disaster (e.g., hurricane-related flooding) to protect emergency workers and public health. A practical method for prioritizing areas for emergency mosquito control has been developed and is described here. North Carolina (NC) One Map was used to identify state-level data layers of interest based on human population distribution and mosquito habitat in Brunswick, Columbus, Onslow, and Robeson Counties in eastern NC. Relevant data layers were included to create mosquito control treatment areas for targeted control and an 18-step protocol for map development is discussed. This protocol is expected to help state, territorial, tribal, and/or local public health officials and associated mosquito control programs efficiently create treatment area maps to improve strategic planning in advance of a disaster. This protocol may be applied to any NC county and beyond, thereby increasing local disaster preparedness.

## INTRODUCTION

Strategic planning prior to a natural disaster, such as a hurricane, is essential for counties to prepare for mosquito issues that will inevitably arise as flood waters recede [1–5]. In North Carolina (NC), agencies such as the NC Department of Public Safety, Division of Emergency Management and NC Department of Health and Human Services (NC DHHS) [Division of Public Health (DPH)], along with others, work together to develop communication and contingency plans for post-hurricane activities related to mosquito control.

Geographic information systems (GIS) are valuable tools that can be used for many purposes, i.e., from straightforward mapping/tracking of mosquito habitats near human populations that could be potential treatment areas to spatiotemporal analyses modeling predictions of disease risk [6–9]. Mapping can also be used for determining placement of sentinel chickens used in surveillance for some arboviruses [10]. Ideally, maps of treatment areas would be created and available pre-disaster to ensure the most streamlined ground or aerial response. This planning step will save organizations time and reduce stress on employees and management during the disaster event and maps can also be used during non-disaster periods. Map layers can help identify areas to help mosquito control operators develop surveillance and treatment plans, including flight patterns [11] and supporting ground operations. Layers within the GIS can also show other types of data such as larvicide/adulticide applications, areas where insecticide resistance has been detected, disease cases, as well as mosquito landing and trap counts pre- and post-treatment for reporting/analysis. Protected areas (e.g., NC Heritage Program [NCHP], United States Fish and Wildlife Service [USFWS], major water bodies [streams, rivers], federal land [e.g., nature preserve, conservation easement]) where treatments should be avoided can also be shown.

Organizations dealing with emergency response and local/state mosquito control programs (MCPs) can use baseline mapping information to develop a customized response for their respective communities. Baseline maps provide an important planning foundation, especially for communities with no mosquito control experience. Government administrators can use the information to estimate costs and readiness for justification of services through the Federal Emergency Management Agency (FEMA) application process post-disaster (i.e., mosquito habitat identification, protected areas of restricted insecticide application). GIS can help MCPs plan locations for larval surveillance, mosquito landing counts, and trapping pre- and post-insecticide application. This will aid MCPs in providing recommendations to local health directors, as well as help with press releases and communication with public information officers. Pre-planning and active communication between agencies involved in emergency response ensures that local management personnel have knowledge of the mosquito response plan in advance of a disaster [3,6,12].

Hurricanes and flooding can increase the potential for mosquito-human contact and incidence of mosquito-borne disease [6,12]; however, this relationship is complex [13]. A federal declaration of a disaster or emergency renders state/local agencies eligible for disaster-related reimbursement [14]. FEMA works with CDC to determine the extent of mosquito-borne disease risk. GIS can be a valuable tool for health directors who (in NC) are the deciding authority on emergency mosquito control response regarding reimbursement rules by FEMA [15]. It would be a good practice to consult local municipalities on whether they would like to be included on an emergency mosquito control response at the county level [3].

Naled is an organophosphate insecticide active ingredient commonly used in aerial application for mosquito control [16]. In 2018, in response to the aftermath of Hurricane Florence in NC, VDCI conducted aerial mosquito control treatments using naled in four counties (Brunswick, Carteret, Harnett, Johnston) encompassing 919,272 acres [11]. Despite *ca*. $3.5 billion of state and federal funding spent on recovery efforts from Hurricanes Matthew (2016) and Florence (2018) [17], there was a lack of preparedness (e.g., lack of updated maps, communication issues) of many eastern NC counties after Hurricane Florence. This lead to the development of a statewide mosquito control services contract (duration of current contract: August 23, 2021-August 24, 2024) with a private vendor who would help manage emergency mosquito control (https://files.nc.gov/ncdps/documents/files/NCDPS-MAC-Fact-Sheet-Oct-20-2021-v1.pdf) through the NC Department of Public Safety, Emergency Management. In 2021, NC created a Mosquito Management Task Force composed of personnel from NC Department of Public Safety, NC DHHS [DPH], county mosquito control, and representatives from the non-profit NC Mosquito and Vector Control Association to assist with the strategic planning and implementation process of an emergency mosquito control response. Currently, no known published studies have described the stepwise process for identifying and mapping mosquito control treatment areas and this could be used for normal and/or emergency planning.

Consequently, the objective here was to develop a straightforward GIS protocol for mapping possible areas to direct aerial and/or ground insecticide treatments for mosquito control post-hurricane. This protocol provides a baseline foundation for organizations that will improve preparedness and reduce stress during the post-disaster period. Ideally, the creation of maps could be standardized and used by any county across NC or other areas using an autonomous analysis (i.e., model builder, Python script), adaptive layers (e.g., population distribution/density), national datasets (e.g., wetlands, flood plains), heritage data (e.g., managed/restricted areas), water bodies, and other layers using state-level data.

## MATERIALS AND METHODS

### Development of Geographic Information System

NC One Map (https://www.nconemap.gov/) was used to identify state-level data layers of interest based on population distribution and mosquito habitat in Brunswick, Columbus, Onslow, and Robeson Counties in eastern NC. A Python script was integrated into the model and customized to the needs of MCPs. Data layers of interest included: human population density that was local address point data originating from the local authority/county/municipality in conjunction with block-level population data from the US Census Bureau [18–20], wetlands [21], county boundaries, protected areas (i.e., NCHP data from USFWS, major water bodies [streams, rivers] [22–23], and federal land [e.g., nature preserve, conservation easement]). A municipal data layer was also added so health directors could do a local evaluation of the treatment zone maps prior to deciding the extent of emergency mosquito control measures.

### United States Fish and Wildlife Service Review

The USFWS must review proposed mosquito control treatment areas for presence of endangered species prior to treatment. This federal agency may recommend changes based on knowledge of endangered species in the area that may be impacted by insecticide application, based on organism biology. Although this review is required, the maps provided here have not yet undergone final review by USFWS.

### Mapping Mosquito Control Treatment Zones

A 1 mi^2^ index grid was developed to demonstrate incidence of address points (built structures) and delineate areas to be treated. The grid is the foundational component for developing treatment zones and is independent from address point data; however, it can be used to show address density. All known protected areas were removed from the grid since they were not to be treated. Warning zones (i.e., areas within 100 ft of protected areas) and mosquito habitats (i.e., wetlands) were identified and map layers created. In NC, different types of wetlands are known habitats for mosquito species such as: tidal salt marsh, *Aedes taeniorhynchus, Ae. sollicitans*; bottomland swamp, *Ae. infirmatus, Ae. atlanticus, Psorophora* spp., *Anopheles crucians, An. quadrimaculatus, Culex nigripalpus, Cx. erraticus, Culiseta melanura, Coquilletidia perturbans*, and *Uranotaenia* spp.; Atlantic white cedar wetland forests, *Ae. canadensis, Ae. triseriatus, Ae. vexans, Cx. pipiens/quinquefasciatus, Cx. restuans, Cx. territans, Cs. melanura* [24]. *Culex pipiens/quinquefasicatus* and *Cx. restuans* are vectors of West Nile virus. *Culiseta melanura* is the enzootic vector of Eastern equine encephalitis virus. Maps were further analyzed by local mosquito control professionals to identify the suitability of remaining areas within the grid for mosquito control (based on personal knowledge of the county). Here, the population density layer was edited to account for vacation homes and other factors that may influence treatment areas in eastern NC. Address points were used rather than blocks since blocks would not account for vacation homes.

### Automated Analysis for Mapping Mosquito Control Treatment Zones

Environmental Systems Research Institute (ESRI; Redlands, CA) ArcMap software was used to analyze the data to aid local and state officials in determining biologically sensitive, safe, and effective zones for mosquito control application. This process uses geospatial tools such as: select, merge, clip, buffer, erase, grid index features, select layer by location, intersect, feature class to feature class, add fields, and calculate fields.

The analysis contains 18 steps to produce a classified fishnet grid of 1 mi^2^ cells indexed around the address point distribution. The classified grid identifies three zones to be considered by local authorities and MCPs: 1) Approved: Treatable area (mosquito habitat), 2) Warning: Treatable area (area encompasses protected areas that were removed/erased from the grid, leaving only treatable areas), and 3) Not significant: Not treated (areas where people live or structure(s) exist but habitat not ideal for concentrated mosquito populations).

*Step 1*. Select the county to be used in the analysis. The current model can be used for statewide analysis; however, due to database size including considering distribution of address points and lengthy processing time, the stepwise example provided here analyzes a single county. The *select* tool in this step, uses structured query language (SQL): “County = ‘X_0_’” where X_0_ is the desired county.

*Step 2:* The data layer from the NC NHP must be edited to remove extraneous data such as logging or industrial information before analysis to help isolate appropriate areas for mosquito control. The SQL expression [GAP_STATUS = X_0_ Or GAP_STATUS = X_1_ Or OWNER_TYPE = X_2_] can be used where X_0_ is ‘1: Managed for biodiversity; disturbance events proceed or mimicked’, X_1_ is ‘2: Managed for biodiversity; disturbance events suppressed’, and X_2_ is ‘3: federal land’.

*Steps 3 and 4*. Step 3 adds a 50 ft line *buffer* to each side of the center line of streams and rivers. Step 4 includes *merging* the polygons for water bodies, buffered streams and rivers, NHP managed and/or biologically sensitive areas (e.g., endangered species) (from Step 2).

*Steps 5-7*. Step 5 *clips* the merged polygon layer (from Step 4) based on the county-specific polygon (from Step 1). The clip tool works as a cookie cutter, trimming layers outside its boundary lines. Step 6 adds a 100 ft line buffer to the merged layer (from Step 5) and creates a single feature. This buffer helps minimize insecticide droplets that could drift into biologically sensitive areas by extending the distance between the release of the insecticide and the sensitive area. Step 7 clips the statewide wetland layer to the county boundary.

*Steps 8-9*. Step 8 uses the *grid index features* with cells set to 1 mi^2^ to generate a fishnet style grid indexed to the address point layer. Step 9 uses the *erase* tool to eliminate cells with biologically sensitive areas (+100 ft buffer). The remaining cells become the base for the resulting treatment zone.

*Steps 10-12*. Step 10 uses the *select by location* tool to select (from the fishnet grid) cells within 100 ft of the biologically sensitive areas (+100 ft buffer). Step 11 creates a new feature class from the selection using the *feature class to feature class* tool. Step 12 calculates the newly created Type field classified as “Warning”.

*Steps 13-16*. Step 13 involves selecting the fishnet grid by location within 100 ft of the biologically sensitive areas (+100 ft buffer) and the selection is inverted. In Step 14, a new feature class is created from the inverted selection (i.e., new Type field classified as “Not Significant”). Step 15 involves selecting cells from the “Not Significant” grid area that intersect with the wetlands layer. Step 16 calculates the newly created Type field classified as “Approved”.

*Step 17*. In Step 17, the *merge* tool is used to combine the “Warning” grid area with the “Approved” or “Not Significant” grid area. This results in a completely classified grid where 1 mi^2^ cells are indexed on the distribution of address points.

*Step 18*. Step 18 uses the *intersect* tool, targets the fishnet grid, and adds 2020 Census Bureau total population information. The final output of this analysis is a classified grid indexed on distribution of buildings and population. This grid can be classified (type field) either by mosquito-related environmental variables or human population if areas near humans are to be targeted with insecticides.

Once all model steps are completed, polygons can be created delineating areas that could be targeted for mosquito control. This can be evaluated by the county health director and/or other individuals before any operational and financial decisions are made about emergency mosquito control response. This protocol develops a foundation and helps organizations understand the scope of work needed for control.

Here, four eastern NC counties (Brunswick, Columbus, Onslow, Robeson) were analyzed and compared. Now that the initial general stepwise process has been developed here, analysis of ground and/or aerial mosquito control treatment zones can be processed in *ca*. 10 min/county. After processing, a digital map is produced and can be used for further deliberation by local authorities.

## RESULTS

Results of this analysis provide local authorities with information needed to develop ground and/or aerial mosquito control treatment zones standardized across the state and develops a scope of work for cost projection based on vetted acres (i.e., total cost of application and amount of formulated insecticide product needed for a specific number of acres).

### Mosquito Control Treatment Zone Maps Without Population Density

Figures 1–4 show treatment zones in Brunswick, Columbus, Onslow, and Robeson Counties created using data layers including wetlands, county boundaries, protected areas, major water bodies, and federal land. A municipal data layer was also added so health directors and/or others could do a local evaluation.

**Figure 1.**
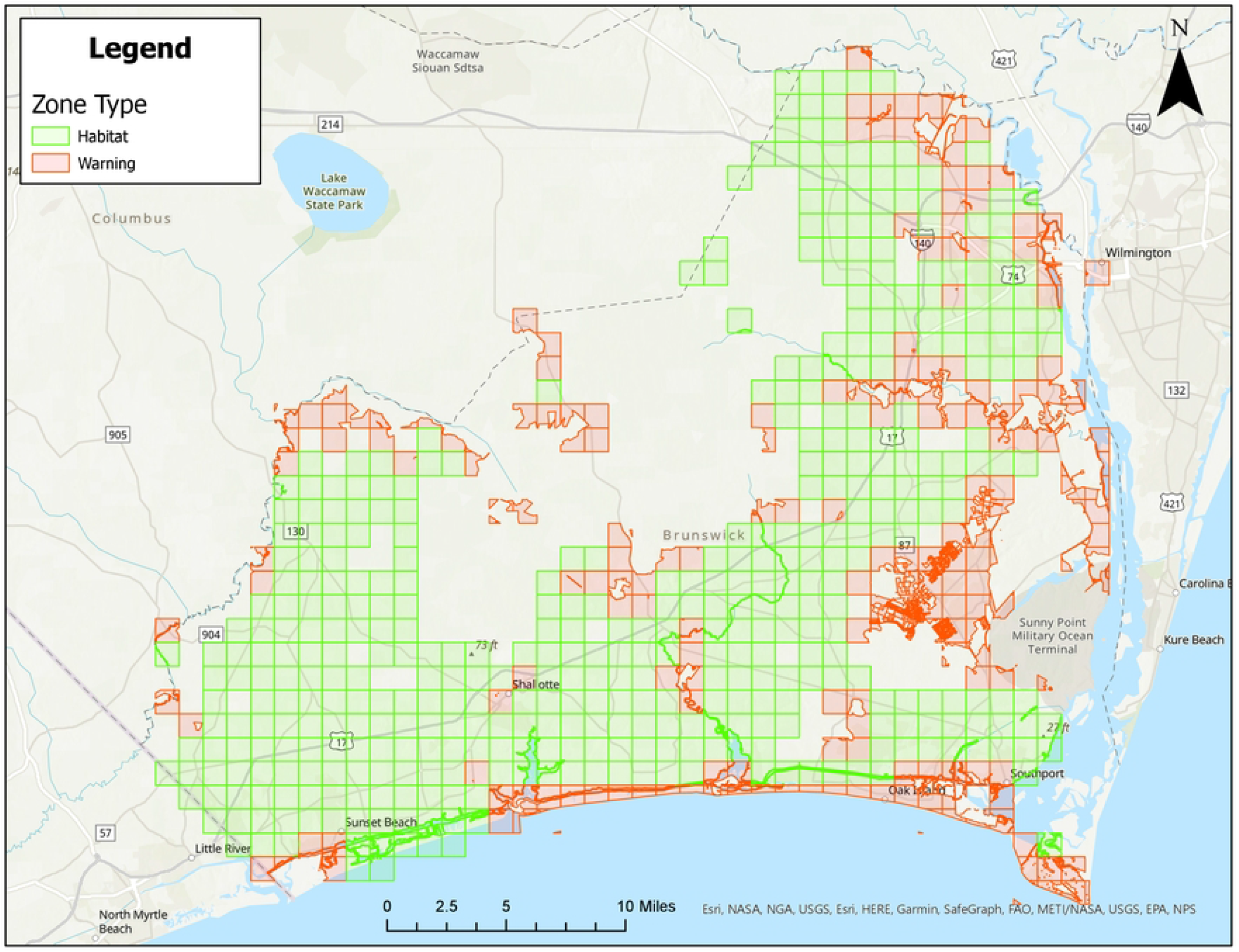
Areas in Brunswick County, North Carolina for treatment consideration classified as approved (green) or warning (red). Maps do not consider human population density.

All counties have areas classified as Approved (green) and Warning (red), but not Non-Significant (due to extensive wetland areas associated with mosquito habitats). Brunswick County’s Approved (369.90 mi^2^) and Warning (212.12 mi^2^) areas total 582.02 mi^2^ classified as areas for treatment consideration (Figure 1). Columbus County’s Approved (640.45 mi^2^) and Warning (66.21 mi^2^) areas total 706.66 mi^2^ classified as areas for treatment consideration (Figure 2). Onslow County’s Approved (244 mi^2^) and Warning (194.94 mi^2^) areas total 438.94 mi^2^ classified as areas for treatment consideration (Figure 3). Onslow County houses the federal lands of Marine Corps Base Camp Lejeune, hence a large area of this county was considered protected (cannot be treated). Robeson County’s Approved (682 mi^2^) and Warning (22 mi^2^) areas total 704 mi^2^ classified as areas for treatment consideration (Figure 4).

**Figure 2.**
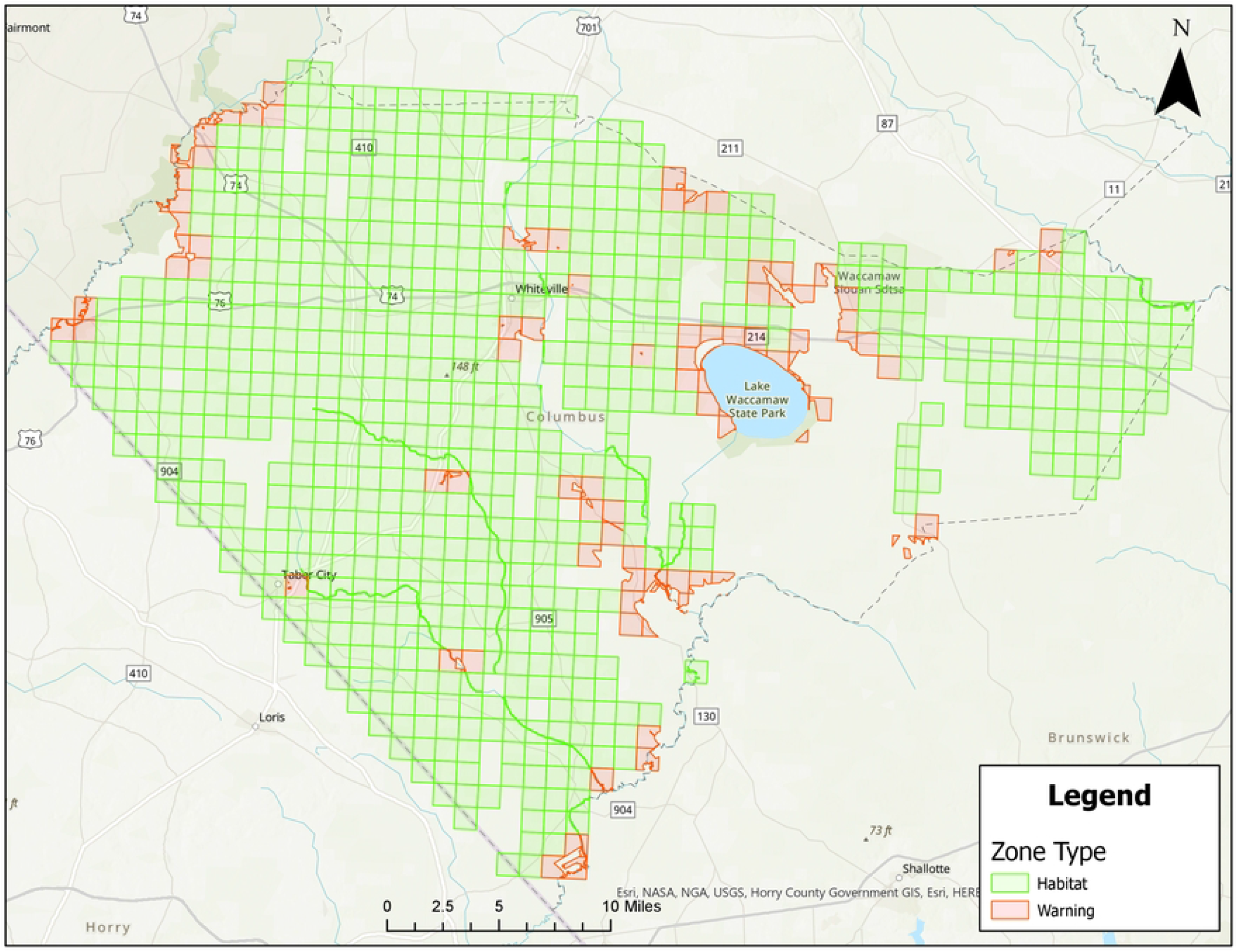
Areas in Columbus County, North Carolina for treatment consideration classified as approved (green) or warning (red). Maps do not consider human population density.

**Figure 3.**
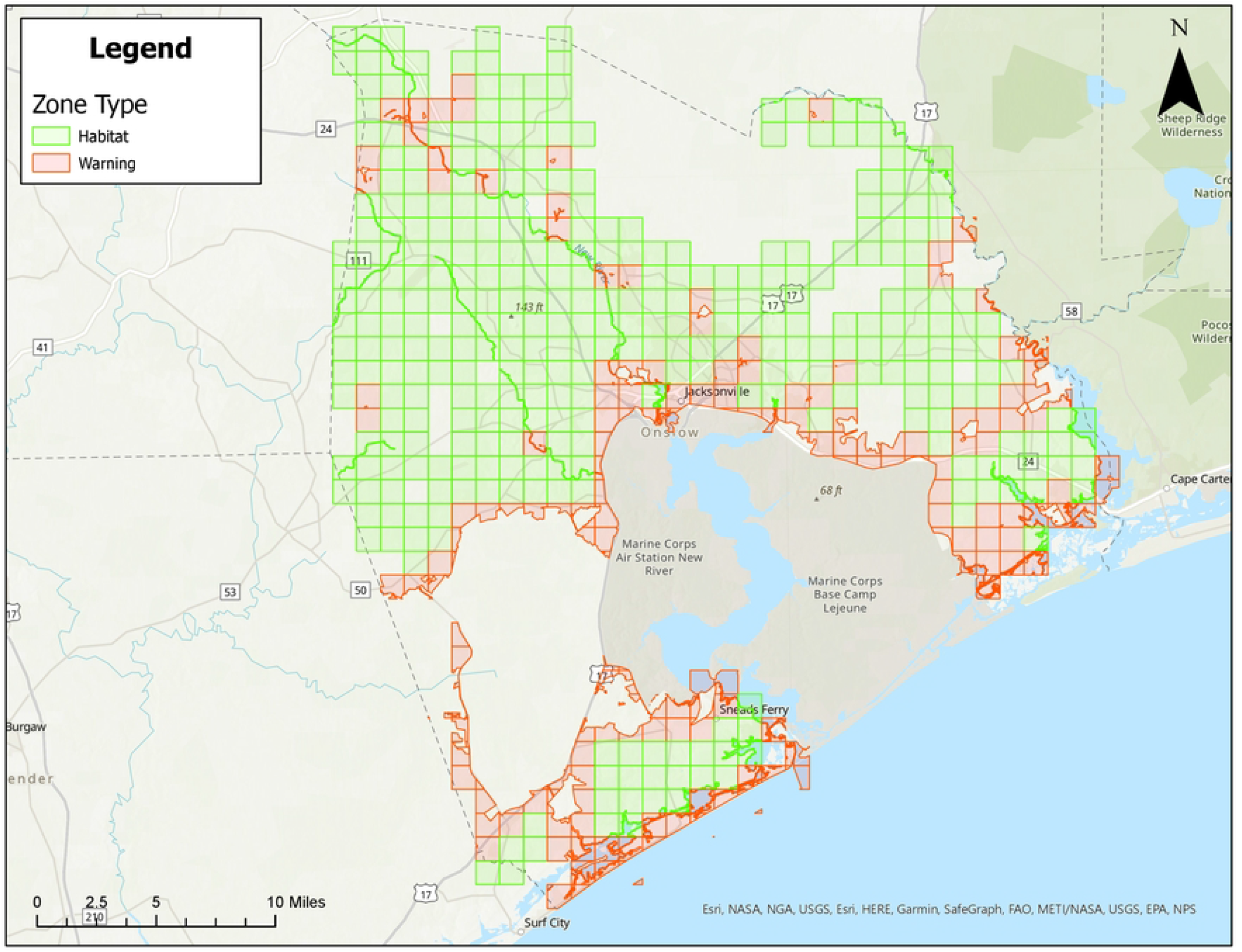
Areas in Onslow County, North Carolina for treatment consideration classified as approved (green) or warning (red). Maps do not consider human population density.

**Figure 4.**
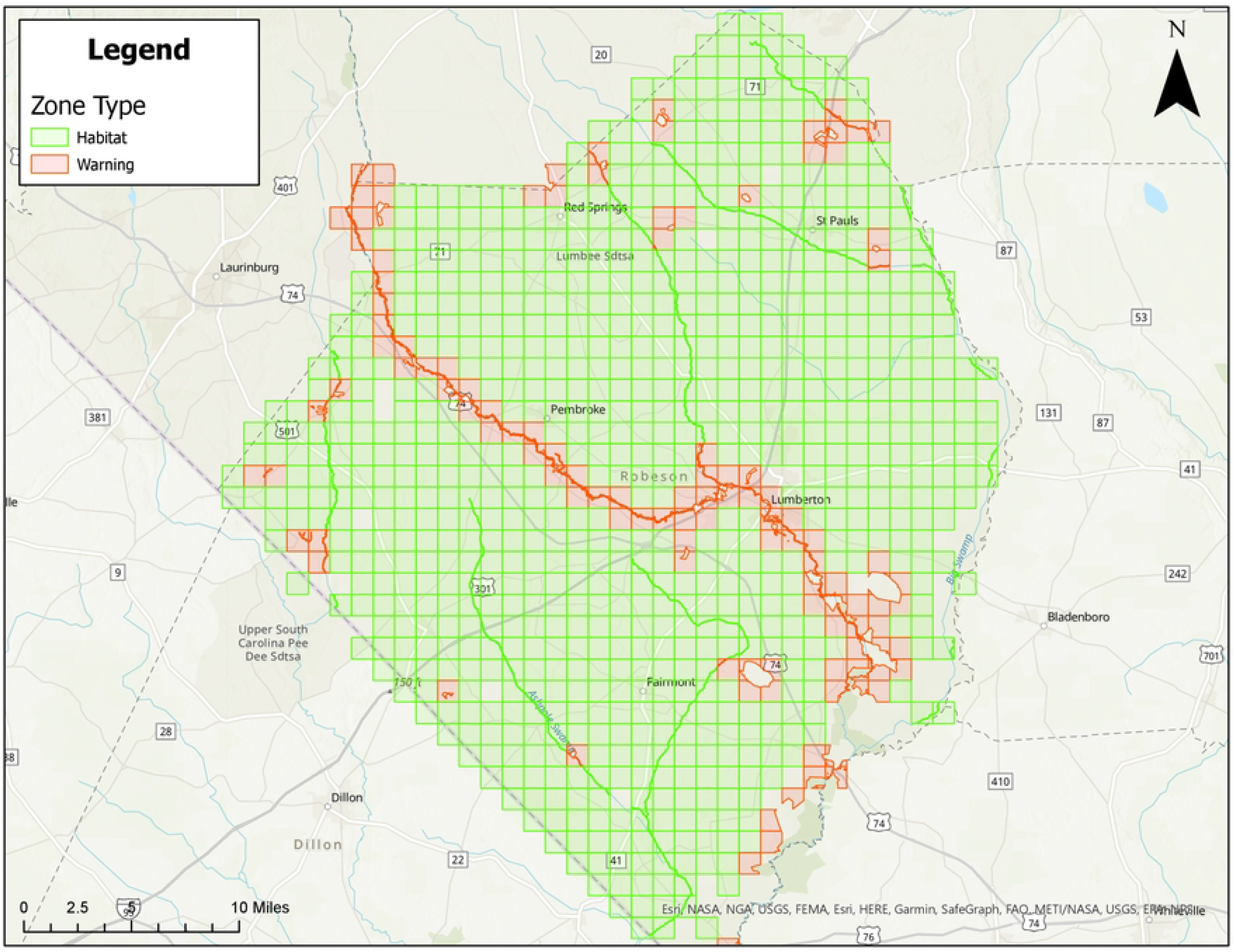
Areas in Robeson County, North Carolina for treatment consideration classified as approved (green) or warning (red). Maps do not consider human population density.

### Creation of Mosquito Control Treatment Zone Maps With Population Density

Figures are also provided showing treatment zones considering human population density in Brunswick (Figure 5), Columbus (Figure 6), Onslow (Figure 7), and Robeson (Figure 8) Counties. Here, people/ft2 is symbolized by colors ranging from white to dark purple. These figures also show the type of classification (i.e., Approved [green] and Warning [red]) data layer overlaid on the population layer. Population maps show concentration of the population in coastal (Brunswick County) and coastal/river/stream (Onslow County) areas, while Columbus and Robeson Counties’ populations are more uniformly spread across the county.

**Figure 5.**
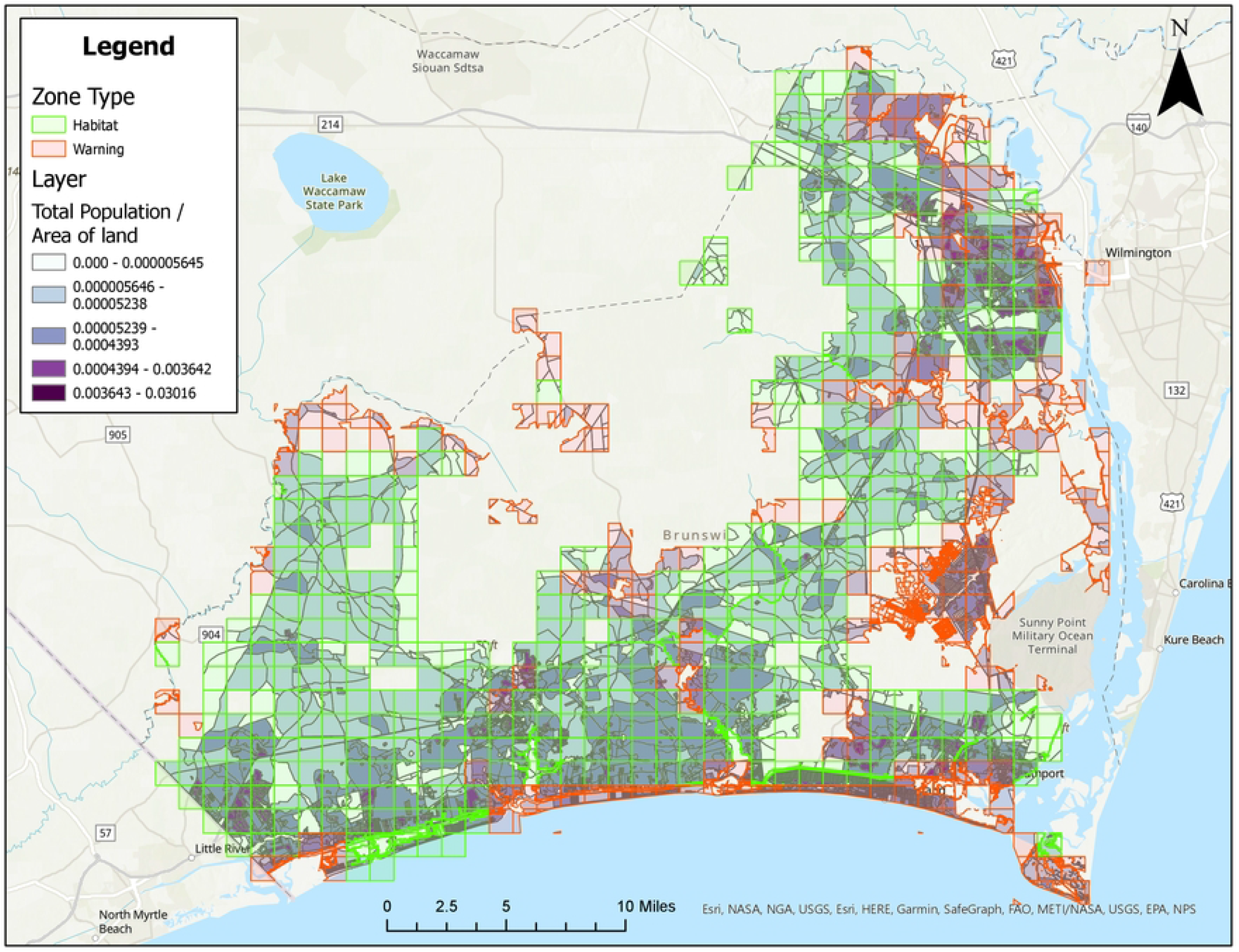
Areas in Brunswick County, North Carolina for treatment consideration classified as approved (green) or warning (red). The human population layer is included and people/ft^2^ is symbolized by colors ranging from white to dark purple.

**Figure 6.**
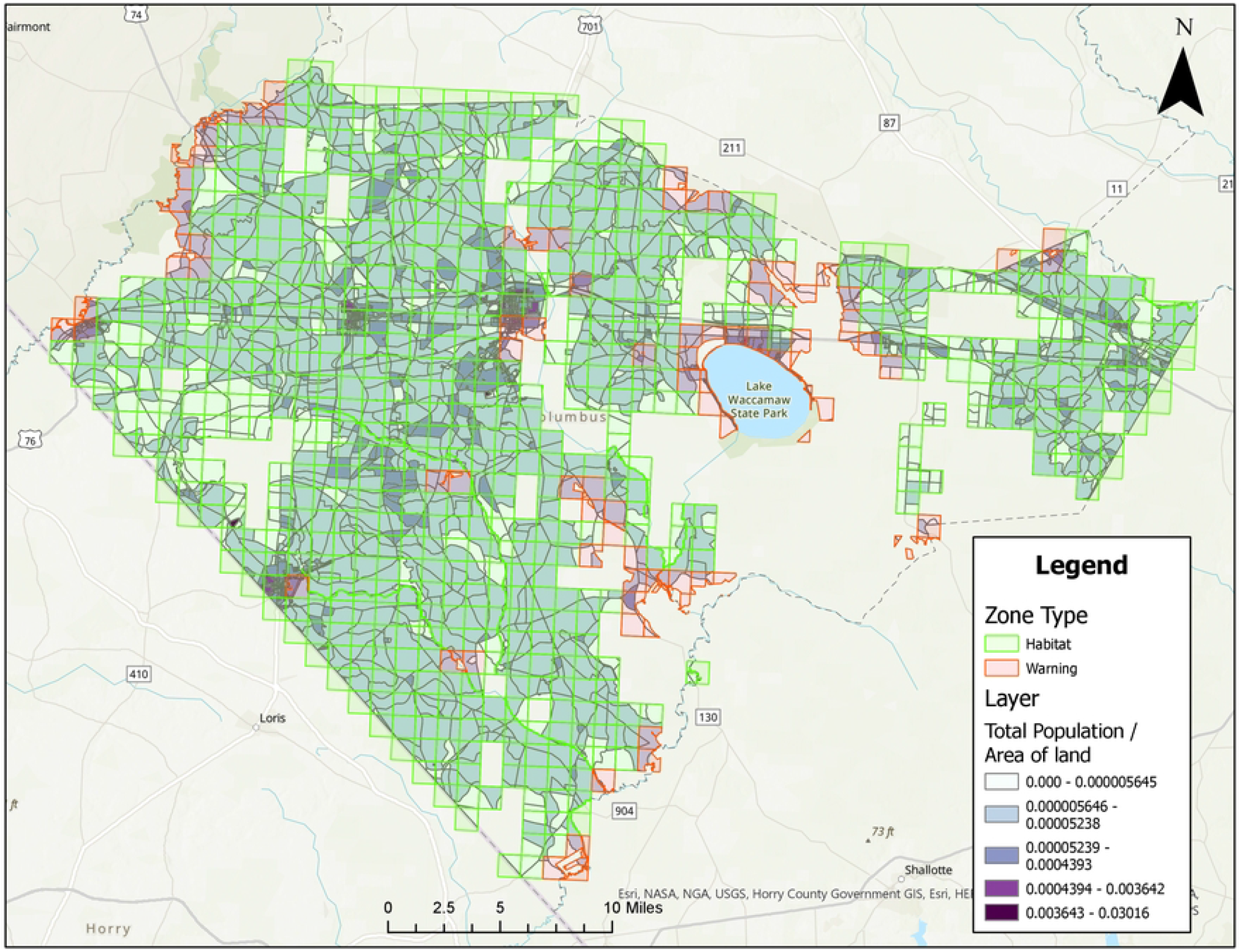
Areas in Columbus County, North Carolina for treatment consideration classified as approved (green) or warning (red). The human population layer is included and people/ft^2^ is symbolized by colors ranging from white to dark purple.

**Figure 7.**
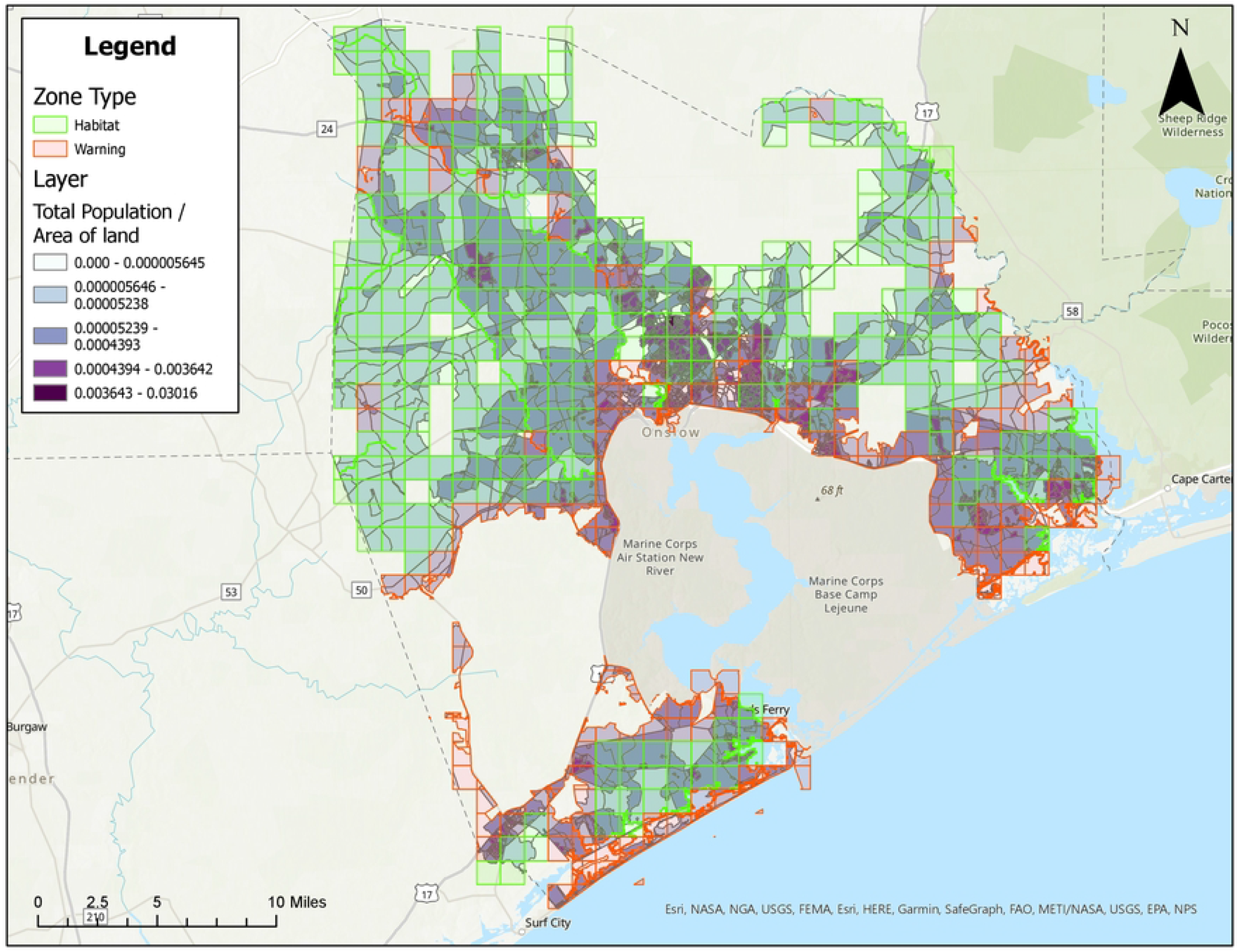
Areas in Onslow County, North Carolina for treatment consideration classified as approved (green) or warning (red). The human population layer is included and people/ft^2^ is symbolized by colors ranging from white to dark purple.

**Figure 8.**
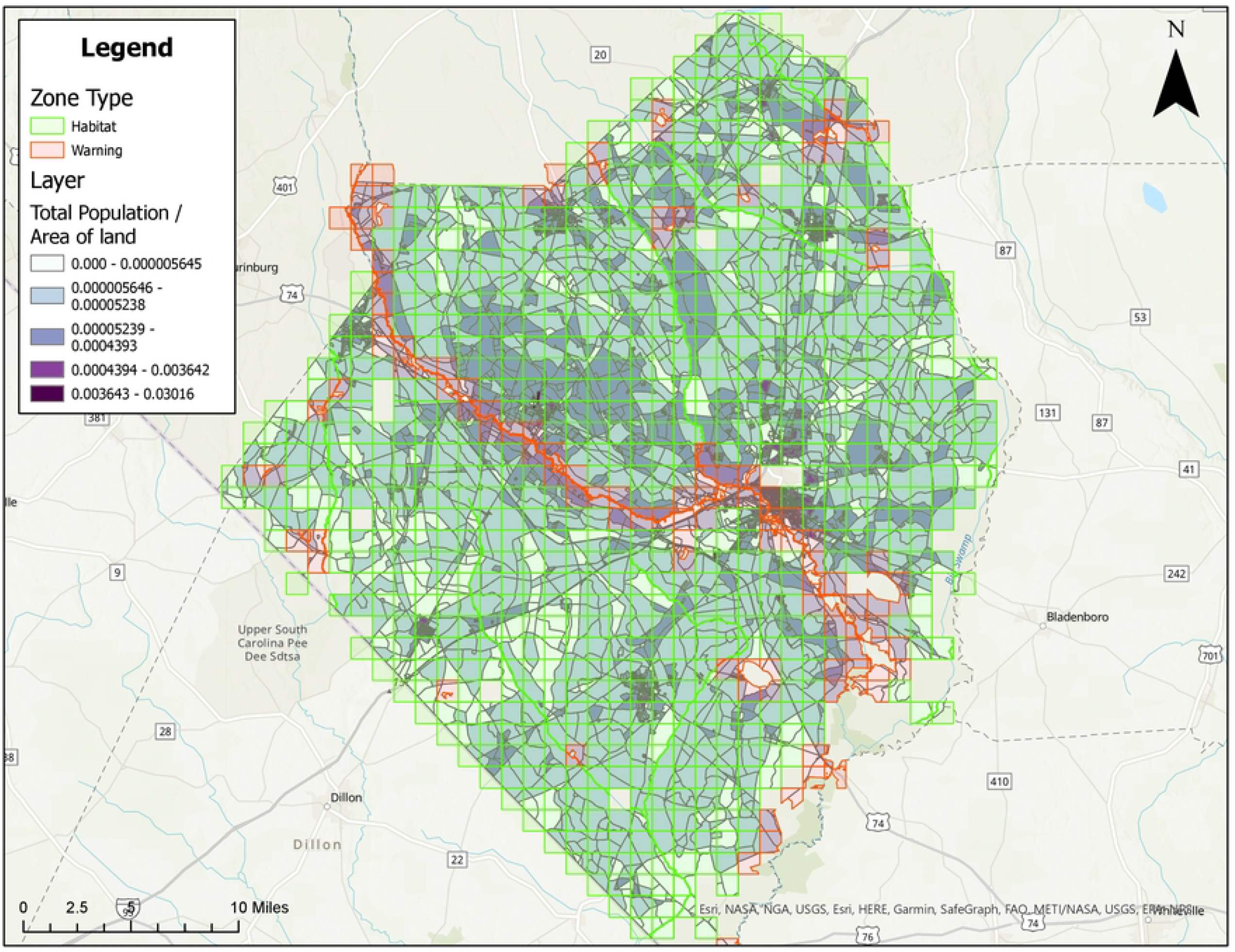
Areas in Robeson County, North Carolina for treatment consideration classified as approved (green) or warning (red). The human population layer is included and people/ft^2^ is symbolized by colors ranging from white to dark purple.

## DISCUSSION

This study provides information on how maps can be created using GIS as part of mosquito control preparedness efforts used in a post-hurricane response. The methodology described here can also be used for normal/routine mosquito control operations. County and/or state GIS or other personnel can use freely available user-friendly data sources to customize map layers for use in mosquito control activities. The NC OneMap resource is shown here, but other states may also have statewide GIS data initiatives that could be utilized (e.g., Florida Geospatial Open Data Portal: https://geodata.floridagio.gov/, Georgia Geospatial Information Office: https://gagiohome-gagio.hub.arcgis.com/). While some mosquito control programs already use GIS for mapping mosquito habitat, the workflow steps and detailed protocol described here are expected to facilitate the planning process of other programs who may be interested in using this type of system.

GIS programs are powerful tools and can streamline mapping and planning processes for mosquito surveillance and control areas, foci of arbovirus outbreaks, and human populations that may be at risk [6–8]. Once counties have these maps with mosquito control treatment acreage delineated, calculations can be made on cost for aerial and/or ground mosquito control so pre-disaster planning can take place between stakeholders. In many cases and with proper planning, the cost of emergency mosquito control response can be partially reimbursed by FEMA [3,15]. The protocol developed here gives local MCPs and other organizations the baseline tools to start working through FEMA requirements in advance of a disaster.

After Hurricane Florence (2018), many NC mosquito control programs were unprepared to deal with mosquito issues related to flooding (e.g., communication issues, lack of emergency response plan), hence more information is needed to improve strategic planning and preparation for these issues that will inevitably occur. This information can be used for both adulticide and larvicide operations [25]. Flood plain and other relevant layers can also be included in this mapping protocol.

Readily available GIS maps will help state, territorial, tribal, local public health officials, and associated local MCPs be well-prepared, reduce stress, and improve the swiftness of any emergency response that may occur. Maps should be updated approximately every five years or as needed to account for changes in land use, landscape ecology, and other factors. Further outreach is necessary to counties in regions where mosquito control response might be needed post-hurricane to improve planning and preparedness activities, including active communication between relevant groups in the chain of command [1,3]. Protocols for GIS development can be shared in advance of a disaster to protect emergency response workers and public health. Baseline maps shown here can be used to demonstrate to health directors and other organizations the areas where mosquitoes are likely to occur post-disaster to help with risk assessments for increased mosquito-human contact. Local authorities such as the health director, county commissioners, and/or MCPs would then determine the areas to be treated and financial decisions can be made.

Studies have shown the need for continued community and financial support for local MCPs to protect health [26–28]. Ideally, local MCPs would be consistently funded to support long-term and well-trained personnel experienced in routine and emergency mosquito control response [27]. Plans should be in place before a disaster occurs to mount an effective response [3,29]. Strategic planning by developing targeted treatment areas in advance of a disaster will help MCPs be better prepared to protect public health.

## Data Availability

All relevant data are within the manuscript and its Supporting Information files.

## ACKNOWLEDGEMENTS

The findings and conclusions in this report are those of the authors and do not necessarily represent the views of the Centers for Disease Control and Prevention.

